# Automatic Sleep Staging from CPAP Airflow using a Dual Fusion Multi-Period Convolutional Neural Network

**DOI:** 10.1101/2025.10.15.25337643

**Authors:** Hsin-Yu Chen, Muneeb Ahsan, Andrey V. Zinchuk, Aatif Husain, Henry K. Yaggi, Hau-Tieng Wu, Cheng-Yao Chen

## Abstract

Background: Continuous Positive Airway Pressure (CPAP) therapy is the standard treatment for obstructive sleep apnea-hypopnea syndrome, yet its use as a passive sleep dynamics monitoring remains limited. CPAP devices record only airflow signals, called CPAP-flow, which constantly interact with the pressure delivered by the device. This interaction renders the signal vulnerable to device-related artifacts and inter-/intra-patient variability, posing significant challenges to its repurposing for monitoring sleep dynamics. Methods: Motivated by neural network-based studies of sleep-wake transition from CPAP-flow, we leverage existing physiological knowledge and introduce a *Dual Fusion Multi-Period Convolutional Neural Network (DFMP-CNN)* model. This deep learning architecture leverages multiple period-specific convolutional kernels and a dual-fusion mechanism to jointly encode known short- and long-range temporal dependencies in CPAP-flow, overcoming limitations of traditional fixed-scale models. Results: Extensive experiments demonstrate that DFMP-CNN achieves state-of-the-art performance in CPAP-based sleep staging. On the Yale dataset, it achieves 78.5% accuracy (Cohen’s *κ* = 0.605), with a best case of *κ* = 0.886; on the Duke dataset, it reaches 73.6% accuracy (*κ* = 0.524), with a best case of *κ* = 0.805. Cross-dataset evaluations confirm the model’s transferability across clinical centers and device types, while feature and fusion ablation studies highlight its robustness. Conclusions: DFMP-CNN provides an unobtrusive approach for sleep monitoring using CPAP devices, providing a dual-purpose platform for therapy and longitudinal assessment. Significance: The robust performance of DFMP-CNN across datasets and device types underscores its potential to improve clinical assessment and optimize therapy management.

## 1 Introduction

Sleep is fundamental to overall health, and sleep disruptions are strongly associated with a wide range of disorders [1]. Accurate characterization of sleep stage progression is critical for the diagnosis and management of sleep-related conditions [2].

Polysomnography (PSG), which integrates multiple physiological signals to achieve high diagnostic accuracy, remains the clinical gold standard for sleep staging.

However, it requires specialized equipment, dedicated facilities, and manual scoring by trained technicians, making it costly, labor-intensive, and impractical for large-scale or longitudinal monitoring. Thus, there is a pressing need for personalized approaches that can conveniently, economically and reliably evaluate sleep dynamics on a daily basis.

Continuous Positive Airway Pressure (CPAP) therapy is the frontline therapy for subjects with Obstructive Sleep Apnea-Hypopnea Syndrome (OSAHS) used nightly by millions in home settings [3]. Modern CPAP devices continuously record airflow signals, enabling fully passive, long-term data collection without patient effort. We refer to this signal as CPAP-flow to distinguish it from the airflow signals recorded by other means. CPAP-flow encodes rich sleep dynamics [4] and has been utilized to detect transitions from sleep to wakefulness using neural network [5]. While it does not provide epoch-by-epoch sleep staging, it has motivated SensAwake, a pressure relief technology used to improving adherence to CPAP devices. SensAwake depends on detecting irregular respiration indicative of wakefulness [6, 7]. On the other hand, several automatic algorithms for epoch-by-epoch sleep staging, including wake, rapid eyeball movement (REM), and non-REM (NREM), have been developed using airflow recorded from PSG [8, 9, 10, 11, 12]. Based on these results, a natural question to ask is if it is possible to develop an automatic epoch-by-epoch sleep staging algorithm using CPAP-flow that is suitable for patients receiving CPAP therapy.

To this end, Figure 1 illustrates how CPAP-flow encodes sleep stage dynamics. Although CPAP therapy introduces interference from the device-delivered pressure, the CPAP-flow signal nonetheless retains meaningful physiological information. For example, wakefulness is characterized by disorganized patterns compared to REM and NREM sleep, with the instantaneous respiratory rate (IRR) remaining relatively stable during NREM. Despite the challenges posed by the absence of additional physiological signals, substantial inter-/intra-patient variability, and susceptibility to device-induced artifacts, these distinct waveform characteristics highlight the potential for a practical, scalable, and personalized alternative for monitoring sleep dynamics directly in the home setting. Supporting this possibility, a recent study [13] demonstrated that topological features of CPAP-flow combined with a boosting-based learning algorithm developed in [11] generalized successfully to a CPAP dataset collected at a single medical center.

**Figure 1.**
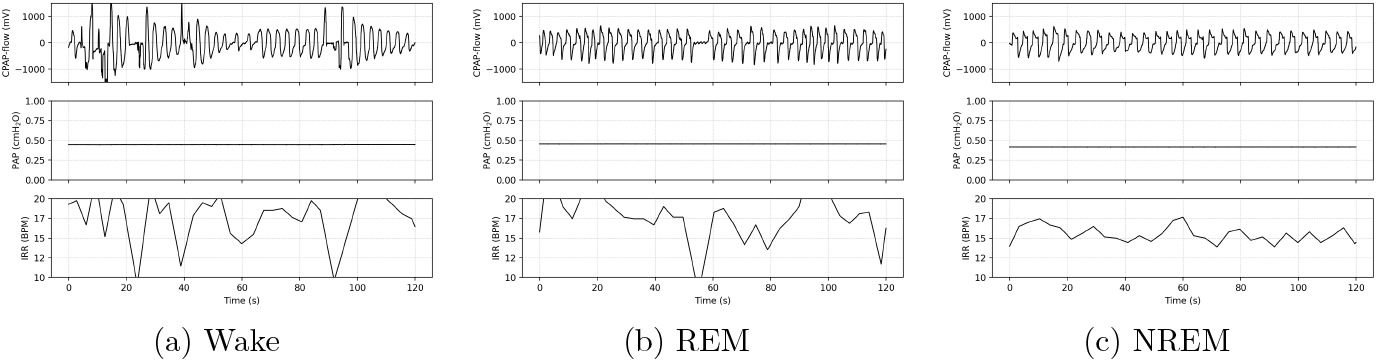
Representative CPAP-flow, PAP and IRR across sleep stages. (a) Wake: irregular CPAP-flow with large fluctuations, yielding variable IRR. (b) REM: smoother rhythm but irregular IRR due to autonomic instability and reduced chemosensitivity. (c) NREM: stable breathing with regular CPAP-flow and smooth IRR. CPAP-flow is expressed in millivolts (*mV*), PAP in centimeters of water (*cmH*_2_*O*), and IRR in breaths per minute (BPM).

Inspired by prior work [5, 13], we hypothesize that advanced modeling strategies specifically tailored to the physiological knowledge associated with sleep and CPAP-flow, and a powerful learning algorithm can enable more accurate monitoring of sleep dynamics. Leveraging established physiological knowledge of multiscale sleep regulation, from cyclic alternating patterns (CAPs) [14] to the interplay among sleep stages, cardiorespiratory coupling, and autonomic nervous system (ANS) activity [15, 16], we propose a novel deep learning framework, the *Dual Fusion Multi-Period Convolutional Neural Network (DFMP-CNN)*. This framework leverages CPAP-flow as a standalone modality to perform automated epoch-by-epoch sleep stage classification (as shown in Figure 2).

**Figure 2.**
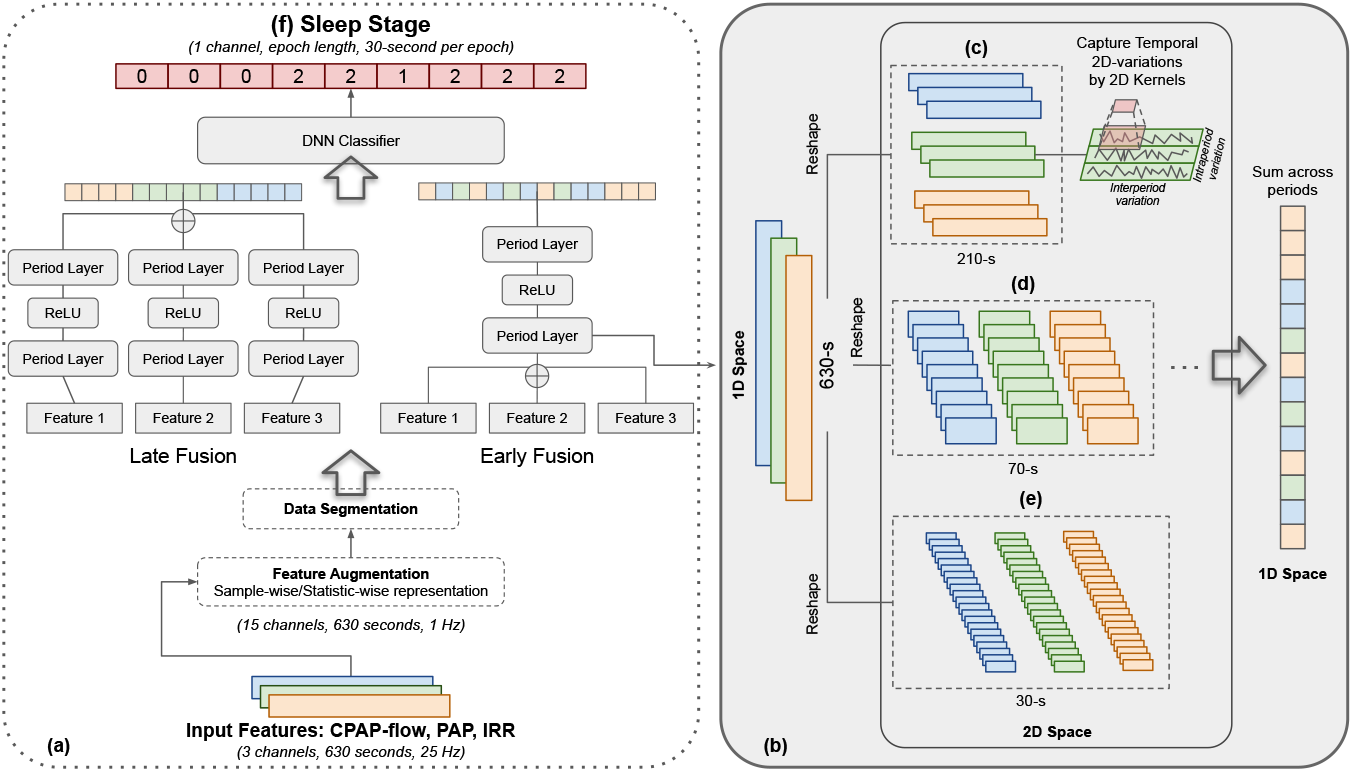
Overall system architecture of the proposed DFMP-CNN model for sleep stage classification using CPAP-derived signals, including CPAP-flow, PAP, and IRR (derived by CPAP-flow). Part (a) presents the overall model structure, while part (b) details the period layer, which reshapes time-series inputs to capture temporal dynamics. Since CPAP-flow and IRR exhibit multi-periodic structures, the model accounts for both intraperiod (within-cycle) and interperiod (across-cycle) variations by transforming one-dimensional sequences into two-dimensional tensors across multiple predefined periods. In our implementation, the input consists of three-channel signals over a 630-second window. Parts (c)–(e) illustrate reshaped feature maps for different periods, e.g., (210 × 3, 3) with a period of 3, (70 × 9, 3) with a period of 9, and (30 × 21, 3) with a period of 21. Finally, part (f) shows the classification output on a 30-second epoch basis, where 0 corresponds to wake, 1 to REM, and 2 to NREM.

The core design of DFMP-CNN lies in its *dual fusion strategy*, which combines early fusion and late fusion to jointly exploit local and global temporal information. Early fusion aggregates features across multiple receptive fields during the convolutional stage, while late fusion integrates predictions from different temporal branches at the decision level, enhancing both robustness and consistency. Here, the convolutional stage refers to the part of the network where convolutional layers extract hierarchical features from the input signal, capturing local temporal patterns at multiple scales before any higher-level integration or decision-making occurs. To further capture the complex, multiscale sleep dynamics presented in respiration, DFMP-CNN integrates multiple period-specific convolutional kernels that operate across diverse timescales. Physiologically, sleep-related respiratory dynamics encompass both short-term fluctuations (e.g., breath-to-breath rhythm changes) and long-term variations (e.g., gradual shifts in respiratory effort and waveform morphology) [4]. A single convolutional kernel with a fixed receptive field is inadequate to represent this wide spectrum of dependencies. Inspired by the multiresolution-based scattering transform [17, 18], which provides a theoretical lens on CNN, DFMP-CNN employs kernels with varying receptive fields, enabling the model to concurrently learn fine-grained micro-patterns and high-level macro-patterns, both essential for accurate and robust sleep stage classification. Furthermore, we leverage how CPAP-delivered pressure impacts CPAP-flow to guide the careful selection of training samples, thereby enhancing the robustness and overall performance of the model.

To evaluate the effectiveness of DFMP-CNN and its generalization, we conducted experiments on two CPAP datasets collected from Yale New Haven Hospital and Duke University Hospital. The proposed DFMP-CNN achieved promising results in three-stage epoch-by-epoch sleep classification, including wake, REM and NREM. In addition to carrying out basic analyses, we conducted further analyses, including zero-shot cross-dataset transfer, demographic subgroup evaluation, and ablation studies on feature and fusion strategies, to validate both robustness and clinical relevance. Despite the inherent black-box nature of deep neural networks, our analysis suggests that the proposed DFMP-CNN can feasibly repurpose CPAP devices as dual-function systems, providing both therapy and passive sleep monitoring.

Moreover, the results demonstrate the model’s transferability across clinical centers, even under variations in devices and acquisition settings.

### 1.1 Related works

Automatic sleep staging has been extensively investigated using diverse signal modalities, particularly home-based systems leveraging reduced-channel. A non-exhaustive set of examples includes electroencephalography (EEG) [19], electrocardigram (ECG) [20], photoplethysmogram (PPG) [21], inertial measurement units (IMU) [22], and multimodal combinations [23, 24], all of which directly reflect parts of physiological processes linked to sleep architecture. In contrast, respiration-only approaches remain relatively underexplored. In addition to some earlier studies [8, 9, 10], Chung et al. [11] extracted topological features from airflow signals but relied on handcrafted representations, limiting adaptability to diverse respiratory patterns. Chen et al.[25] employed respiratory inductance plethysmography (RIP) belt signals, which require additional sensors and offer limited robustness in long-range temporal modeling. Aggarwal et al. [26] modeled nasal airflow using a chain-structured conditional random field (CRF), but such sequential models are constrained in capturing multiscale dependencies. To our knowledge, no prior study has directly leveraged CPAP-flow for sleep staging, except for some prior exploration [5, 6, 7] focusing on detecting the transition from sleep to wakefulness and a recent topological data analysis-based effort [13].

## 2 Methods

The problem of sleep stage classification based on CPAP-flow is a supervised multi-class classification problem. We aim to design a CNN-based algorithm that utilizes temporal dependencies and hierarchical representations to robustly differentiate sleep stages even under noise or missing data. Overall, our proposed framework converts raw CPAP-flow into reliable sleep stage predictions through a three-stage pipeline, as illustrated in Figure 2. First, we extract features directly from the CPAP signal channels, including CPAP-flow and positive airway pressure (PAP), and derive the instantaneous respiratory rate (IRR) to capture multi-scale respiratory dynamics while minimizing redundancy. Second, the extracted features are processed by the Dual Fusion Multi-Period Convolutional Neural Network (DFMP-CNN). In this stage, input signals within a 630-second window are reshaped into structured 2D representations across multiple predefined periods, enabling the model to capture both intraperiod (within-cycle) and interperiod (across-cycle) variations. Finally, the fused representations are passed to a deep neural classifier that predicts sleep stages on a 30-second epoch basis (Figure 2 (f)). We focus on the three-stage setting and denote Y = {0 : wake, 1 : REM, 2 : NREM}.

### 2.1 Feature Extraction

We now detail the feature extraction step.

#### Preprocessing of CPAP-flow

The CPAP-flow is sampled at 25 Hz. The CPAP-flow signal is first detrended by mean removal, then bandpass filtered with a second-order Butterworth filter (0.1–0.35 Hz) to isolate respiratory rhythms while suppressing low-frequency drift and high-frequency noise. A secondary detrending step further stabilizes the signal.

#### Derivation of IRR

Breath intervals are estimated via peak detection [27], identifying local maxima corresponding to exhalation onsets. The time difference between successive peaks yields the respiratory interval, defined as Δ*t*_*i*_ = *t*_*i*_− *t*_*i*−1_ where {*t*_1_, *t*_2_, …, *t*_*n*_} denote exhalation onset times. The set 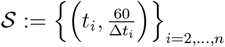 is a nonuniform sampling of the underlying IRR. We interpolate using a monotonic cubic spline to obtain an estimate of IRR, and sample it at 25Hz.

#### PAP

The PAP is sampled at 25 Hz, which records how much pressure CPAP offers over time, which is usually smoother and less dynamic. It potentially encodes transitions between sleep states. We incorporate the raw PAP alongside CPAP-flow and IRR to enrich contextual representation.

#### Normalization

To minimize inter-subject variability and enhance generalization, Z-score normalization was applied independently to each subject across the entire night for every feature.

#### Signal Integrity Mask

CPAP therapy, while effective, introduces unique signal artifacts such as leakage from mask misfit or patient movement, and device disconnections or sensor dropout [28]. To safeguard data quality, we construct a signal integrity mask using CPAP-flow and PAP to flag segments corrupted by leakage, dropouts, or anomalies. Note that pressure jumps due to titration lead to CPAP-flow perturbation. Therefore, for each training segment, we compute the proportion of time points for which *P* ^′^(*t*) ≈ 0. If this proportion exceeds 0.5, the segment is excluded from training to avoid using perturbed data, ensuring the model learns only from physiologically valid, artifact-free signals.

#### Feature Augmentation

Recognizing that sleep dynamics evolve gradually, typically over 30-second epochs, we downsample the original 25 Hz signals to 1 Hz by applying two complementary augmentation strategies to reduce redundancy and enrich the representation [29]:

- *Sample-wise representation*: Every 25 consecutive samples are subsampled, retaining the last value, resulting in an effective sampling rate of 1 Hz [30].
- *Statistic-wise representation*: For each 25-sample window, four descriptors are calculated, including mean, standard deviation, maximum, and minimum, to capture local characteristics [31].

This process yields in total 3×5 = 15 feature channels, sampled at 1 Hz, that balances temporal resolution with descriptive richness.

### 2.2 Model Architecture

To capture the multi-scale temporal structure of respiratory signals, we design the DFMP-CNN, a specialized 2D convolutional framework inspired by TimesNet [32] and related work [33]. Unlike conventional 1D CNNs [34] that process sequences linearly, DFMP-CNN reshapes time series into structured 2D representations through periodic segmentation, enabling richer feature extraction.

#### 2.2.1 Dual-Fusion Strategy

A central design of DFMP-CNN is the *dual-fusion mechanism* [35, 36], which integrates both early and late fusion to balance modality interaction and preservation. *Early fusion* merges low-level features across modalities, allowing initial cross-modal interactions but risking the loss of modality-specific signatures. *Late fusion*, in contrast, processes each modality separately, maintaining distinct statistical and temporal patterns but limiting interdependency capture.

Our approach combines both: CPAP-flow, PAP, and IRR are first processed independently to preserve modality-specific structure. Their outputs are then concatenated into a unified feature map, enabling deeper layers to learn cross-modal dependencies. Early and late fusion are performed in parallel and subsequently combined before being passed to the fully connected layers; thus, their relative order does not influence the results. This dual-fusion design enhances the representation of both intra- and inter-modality dynamics, improving robustness against noise, leakage, and subject variability. See Figure 2 (a) for details.

#### 2.2.2 Multi-Period Convolutional Backbone

On top of the fused representation, DFMP-CNN applies a multi-period 2D convolutional backbone. For each period *p*, an input segment of *T* = 630 seconds (10.5 minutes) is reshaped into a matrix 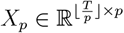, where is the floor function, so that convolutional filters can jointly capture intraperiod patterns (within *p*) and interperiod transitions (across *T/p*). We consider three periods ∈ { *p* 3, 9, } 21 in DFMP-CNN.

This design is informed by well-known multiscale physiological facts. The 630-second context is motivated by capturing microstructural rhythms in sleep such as autonomic cycles and CAPs [37, 14]. Each selects *p* reflecting a distinct scale of sleep dynamics. *p* = 3 captures fine-scale respiratory cycles and short-term dynamics of ANS [38, 39]. *p* = 9 captures mid-scale autonomic arousals and EEG micro-bursts [40, 41]. *p* = 21 captures longer dynamics such as K-complex clusters and CAP phases [42, 43].

The multi-period convolutional features are passed through layers with kernel size 3 (i.e., covering 3 consecutive samples) and ReLU activation [44, 45]. Different periods, each corresponding to a distinct temporal reshaping of the input, are used to generate 2D representations that are individually convolved with kernels. The resulting feature maps from all periods are then reshaped back to 1D and aggregated by summation across the period dimension, thereby integrating information from multiple temporal scales while retaining intra-period characteristics. This aggregated representation is subsequently projected through fully connected layers to yield the final sleep stage classification scores. The projection maps the high-dimensional, multi-period convolutional features into the label space, enabling the model to capture cross-period dependencies and deliver robust sleep stage predictions (see Figure 2 (b) for an illustration).

### 2.3 Implementation

We describe the implementation steps of our model to enable efficient training.

#### 2.3.1 Data Segmentation

Overnight signals from all 15 channels were segmented using a rolling window approach with a step size of 2 data points. Each input segment had a duration of *T* = 630 seconds, yielding a 2D representation of size *N*_segments_ × 630 as a *training sample*, where *N*_segments_ is the total number of generated segments. A signal integrity mask was then applied to exclude low-quality segments, and the remaining high-quality segments were used as model inputs for training.

#### 2.3.2 Ground-Truth Label Assignment

For each training sample, the corresponding ground-truth label is assigned as follows. Each 630-second (10.5-minute) segment is first divided into 21 consecutive 30-second epochs, which aligns with the expert annotations provided at the standard 30-second resolution for sleep staging. A majority voting scheme is then applied across these 21 epochs: the label that appears most frequently within the segment is assigned as the representative label for the entire 10.5-minute segment. To ensure label reliability, we calculate the dominance ratio of this label, defined as its proportion among the 21 epochs. Only segments with a dominance ratio of 1.0 (i.e., all 21 epochs have the same label) are retained for training; segments with any disagreement are discarded. This procedure ensures that each training segment is associated with a consistent and representative label, thereby reducing the risk of noisy supervision from transient stage transitions.

#### 2.3.3 Model Settings

The model is trained with a weighted cross-entropy loss [46] to address class imbalance across sleep stages. Optimization is performed using Adam [47] with an initial learning rate of 1 × 10^−4^, decayed when the validation loss plateaus, and early stopping is applied to prevent overfitting. Regularization techniques include dropout (*p* = 0.5), *L*_2_ weight decay (1 × 10^−4^), and batch normalization [48]. To enhance robustness, we apply mild signal augmentations such as temporal jittering [49] and amplitude scaling, simulating realistic breathing variability. The model is implemented in PyTorch and trained on NVIDIA A100 GPUs, with each experiment repeated across multiple random seeds for reproducibility.

## 3 Material

This section describes the materials, including the datasets, the baseline models for comparison, and the evaluation protocol used in the experiments.

### 3.1 Datasets

This study draws on overnight CPAP titration recordings collected at the sleep centers of Yale New Haven Hospital and Duke University Hospital. All signals were recorded directly from CPAP devices at 25 Hz and paired with sleep stage labels manually scored by certified technicians according to AASM guidelines [50].

The Yale New Haven Hospital (abbreviated Yale afterward) database comprises 100 subjects previously diagnosed with OSAHS, who underwent in-lab attended positive airway pressure titration PSG at the Yale Sleep Center between January 1, 2018, and September 1, 2023. Those subjects were monitored during in-lab PSG with concurrent CPAP titration using ResMed S9 VPAP Tx and Philips Respironics OmniLab Advanced Tx units. Participants ranged from 19 to 90 years of age, with recording sessions lasting at least 5 hours. Within this cohort, seven subjects had no recorded REM sleep and 34 lacked deep sleep stages. The Duke University Hospital (abbreviated Duke afterward) database includes 36 subjects who underwent split-night PSG and CPAP titration on a ResMed S9 VPAP Tx device. All participants were at least 20 years old, with suspected OSAHS confirmed during the study. Recordings lasted at least 2.5 hours (average 4.2 hours). Two subjects exhibited no REM sleep, while 21 showed no deep sleep stages.

In the Yale dataset, 8 out of 100 recordings were excluded due to poor data quality, primarily from transmission errors or sensor disconnections, leaving 92 recordings for analysis. Participants exhibited a mean total sleep time (TST) of 363.2 ± 72.2 minutes, comprising 62.3± 43.1 minutes in REM sleep and 300.6 ± 64.6 minutes in NREM sleep. In the Duke dataset, 4 of 36 recordings were excluded, one for to TST less than 60 minutes and three for poor data quality, yielding 32 recordings for analysis. Participants had a mean TST of 197.9 ± 54.6 minutes, with 46.5 ± 23.4 minutes in REM and 151.1 ± 43.9 minutes in NREM.

## 3.2 Benchmark

Since CPAP-flow is a relatively novel input modality for sleep stage classification, we establish fair and comprehensive comparisons against representative state-of-the-art (SOTA) baselines by reproducing prior works and adapting their methodologies to our dataset.

- *Conventional ML Models*. Prior studies such as Chung et al. [11] and Chen et al. [25] extracted handcrafted descriptors utilizing toplogical data analysis and employed boosting-based classifiers for sleep staging. Following this line, we implemented (i) XGBoost, (ii) LightGBM, and (iii) Random Forest to assess the performance of feature-driven approaches.
- *Deep Learning Models*. Wei et al. [51] demonstrated that deep architectures could achieve strong performance for coarse-grained sleep classification. Inspired by their framework, we implemented two neural baselines adapted to

CPAP-flow: (iv) a Dnn, which flattens the input sequence and passes it through fully connected layers to capture global temporal dependencies; and (v) a Cnn, which applies convolutional filters to extract localized temporal dynamics before feeding the representations into dense layers for classification. The hybrid design allows better modeling of both short- and long-term patterns compared to a pure DNN.

### 3.3 Evaluation Protocol

We adopted a three-fold cross-validation scheme on the subject level, where each fold served once as the test set while the remaining subjects were used for training.

Performance was quantified using *accuracy, Cohen’s kappa κ*, and *macro F1-score*, capturing overall correctness, agreement beyond chance, and balanced precision–recall trade-offs under class imbalance. In addition, class-wise measures including *sensitivity* (recall), *positive predictive value* (PPV, precision), and *specificity* were reported to evaluate detection reliability across all sleep stages, with emphasis on underrepresented stages such as REM. To assess alignment with clinically relevant sleep architecture, we computed the *Pearson correlation coefficient (PCC)* between predicted and ground-truth TST, defined as the sum of REM and NREM durations.

## 4 Results

This section shows results of various experiments.

### 4.1 Overall Performance

As shown in Table 1, DFMP-CNN achieves the highest overall accuracy, *κ*, and macro F1 across both datasets, outperforming all reproduced baselines. On the Yale dataset, DFMP-CNN reached an accuracy of 78.5% and *κ* = 0.605, surpassing the strongest baseline (CNN-DNN, *κ* = 0.587). Gains were most pronounced in REM detection, where sensitivity and PPV improved to 0.604 and 0.558, compared with 0.564/0.530 for CNN-DNN and substantially lower values for tree-based methods (e.g., REM sensitivity 0.326 with RF). Importantly, wake detection remained robust, with sensitivity of 0.748 and PPV of 0.774, highlighting the model’s ability to capture arousals and wake–sleep transitions. Specificity across all classes exceeded 0.92, indicating low false positive rates and strong reliability.

**Table 1.**
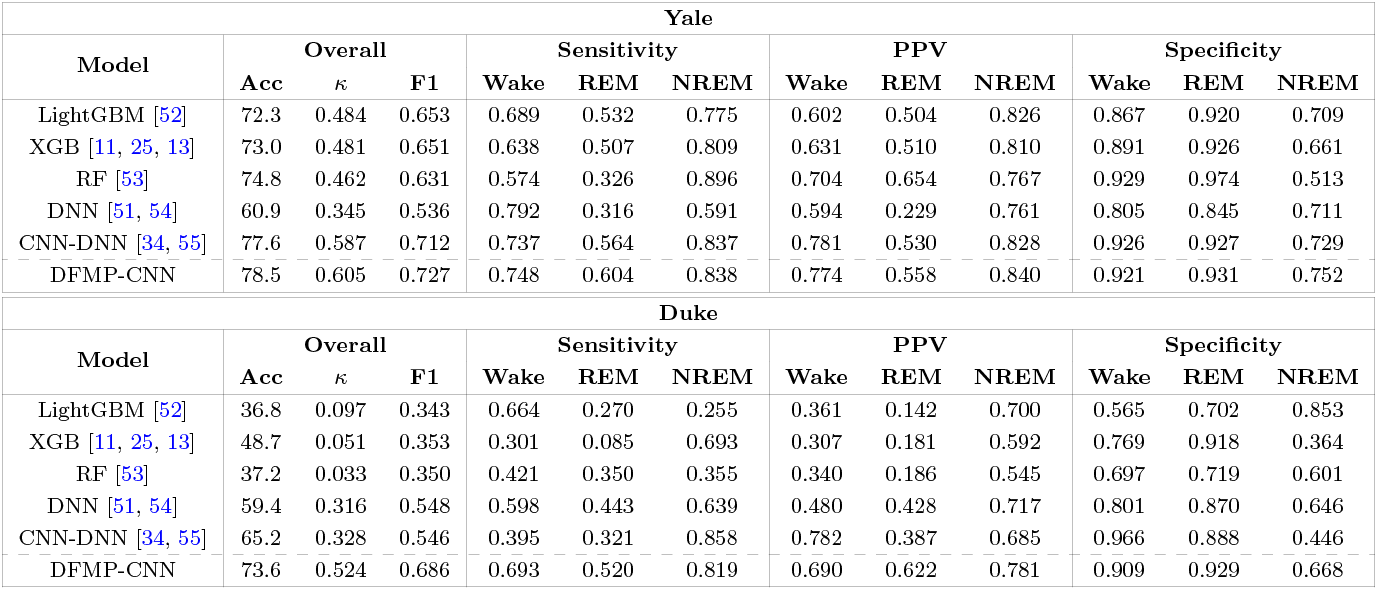
The overall results of 3-stage classifications on Yale and Duke datasets compared with the baselines. Acc: Accuracy; *κ*: Cohen’s kappa; F1: Macro F1-score.

| Yale |  |  |  |  |  |  |  |  |  |  |  |  |
| --- | --- | --- | --- | --- | --- | --- | --- | --- | --- | --- | --- | --- |
| Model | Overall |  |  | Sensitivity |  |  | PPV |  |  | Specificity |  |  |
| | Acc | $\kappa$ | F1 | Wake | REM | NREM | Wake | REM | NREM | Wake | REM | NREM |
| LightGBM [52] | 72.3 | 0.484 | 0.653 | 0.689 | 0.532 | 0.775 | 0.602 | 0.504 | 0.826 | 0.867 | 0.920 | 0.709 |
| XGB [11, 25, 13] | 73.0 | 0.481 | 0.651 | 0.638 | 0.507 | 0.809 | 0.631 | 0.510 | 0.810 | 0.891 | 0.926 | 0.661 |
| RF [53] | 74.8 | 0.462 | 0.631 | 0.574 | 0.326 | 0.896 | 0.704 | 0.654 | 0.767 | 0.929 | 0.974 | 0.513 |
| DNN [51, 54] | 60.9 | 0.345 | 0.536 | 0.792 | 0.316 | 0.591 | 0.594 | 0.229 | 0.761 | 0.805 | 0.845 | 0.711 |
| CNN-DNN [34, 55] | 77.6 | 0.587 | 0.712 | 0.737 | 0.564 | 0.837 | 0.781 | 0.530 | 0.828 | 0.926 | 0.927 | 0.729 |
| DFMP-CNN | 78.5 | 0.605 | 0.727 | 0.748 | 0.604 | 0.838 | 0.774 | 0.558 | 0.840 | 0.921 | 0.931 | 0.752 |

Table 1: The overall results of 3-stage classifications on Yale and Duke datasets compared with the baselines. Acc: Accuracy; $\kappa$ : Cohen’s kappa; F1: Macro F1-score.
| Duke |  |  |  |  |  |  |  |  |  |  |  |  |
| --- | --- | --- | --- | --- | --- | --- | --- | --- | --- | --- | --- | --- |
| Model | Overall |  |  | Sensitivity |  |  | PPV |  |  | Specificity |  |  |
| | Acc | $\kappa$ | F1 | Wake | REM | NREM | Wake | REM | NREM | Wake | REM | NREM |
| LightGBM [52] | 36.8 | 0.097 | 0.343 | 0.664 | 0.270 | 0.255 | 0.361 | 0.142 | 0.700 | 0.565 | 0.702 | 0.853 |
| XGB [11, 25, 13] | 48.7 | 0.051 | 0.353 | 0.301 | 0.085 | 0.693 | 0.307 | 0.181 | 0.592 | 0.769 | 0.918 | 0.364 |
| RF [53] | 37.2 | 0.033 | 0.350 | 0.421 | 0.350 | 0.355 | 0.340 | 0.186 | 0.545 | 0.697 | 0.719 | 0.601 |
| DNN [51, 54] | 59.4 | 0.316 | 0.548 | 0.598 | 0.443 | 0.639 | 0.480 | 0.428 | 0.717 | 0.801 | 0.870 | 0.646 |
| CNN-DNN [34, 55] | 65.2 | 0.328 | 0.546 | 0.395 | 0.321 | 0.858 | 0.782 | 0.387 | 0.685 | 0.966 | 0.888 | 0.446 |
| DFMP-CNN | 73.6 | 0.524 | 0.686 | 0.693 | 0.520 | 0.819 | 0.690 | 0.622 | 0.781 | 0.909 | 0.929 | 0.668 |

Performance on the Duke dataset, which is inherently more challenging due to CPAP titration protocols and a population restricted to moderate-to-severe OSAHS, followed a similar trend. DFMP-CNN achieved 73.6% accuracy and *κ* = 0.524, marking a substantial improvement over CNN-DNN (*κ* = 0.328) and DNN (*κ* = 0.316). Notably, REM detection was significantly stronger with DFMP-CNN (sensitivity/PPV = 0.520/0.622) compared to CNN-DNN (sensitivity/PPV = 0.321/0.387), a relative improvement of nearly 60%. Even under disrupted sleep conditions with frequent PAP adjustments, the confusion matrix in Figure 3 (b) further confirms this balanced performance, showing relatively low cross-stage misclassification rates even in noisy titration conditions: wake sensitivity 0.693, NREM sensitivity 0.819, and corresponding PPVs above 0.68. This demonstrates that DFMP-CNN generalizes well to noisy, clinically adverse conditions.

**Figure 3.**
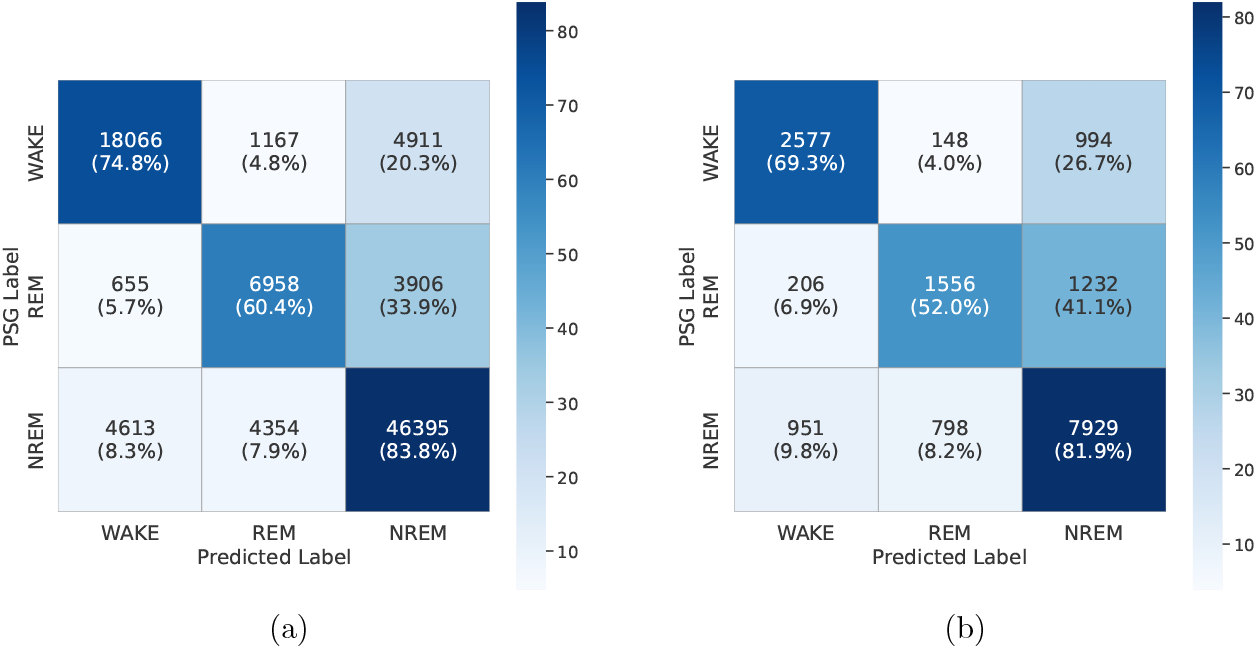
Confusion matrices for the (a) Yale and (b) Duke datasets.

### 4.2 Total Sleep Time Measurement

To verify alignment with clinical sleep architecture, we compared predicted and ground-truth TST. As shown in Figure 4, the model achieved Pearson correlation coefficients (PCC) exceeding 0.7 on both datasets, with regression slopes close to unity and mean biases close to 5 minutes. These results demonstrate that DFMP-CNN not only delivers accurate epoch-wise classification but also preserves global measures of consolidated sleep.

**Figure 4.**
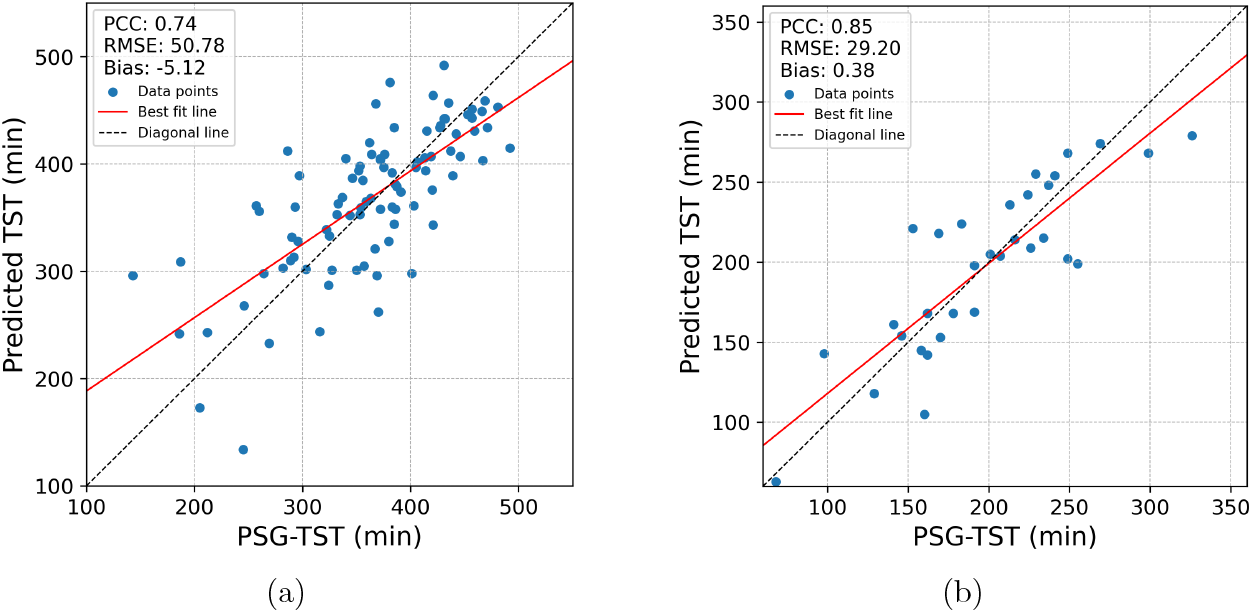
Scatter plot comparing predicted TST to PSG-TST on (a) Yale and (b) Duke datasets. Red solid lines are the least-squares linear regression lines (best fit lines) fitted to the data to illustrate the linear relationship between predicted and PSG TSTs. PCC: Pearson correlation coefficient; RMSE: Root Mean Square Error (min: minute).

### 4.3 Visualization

We present representative overnight hypnograms in Figure 5 to illustrate model behavior. Each panel shows, from top to bottom: CPAP-flow, PAP, and IRR signals, PSG sleep stage labels, predicted sleep stage labels, and model output probabilities for REM (blue) and NREM (orange), with epochs classified as wake when *P*_REM_ + *P*_NREM_ *<* 0.5. The predicted hypnograms exhibit strong alignment with PSG annotations, with subject-level *κ* scores indicating substantial agreement and a clear similarity in the temporal progression of stage transitions. As shown in the zoomed-in Figure 6, REM periods are generally well captured, though some episodes remain underdetected, particularly in early-night cycles when REM is sparse and easily confused with wakefulness. This finding is likely due to their subtle and ambiguous respiratory patterns. Unlike EEG, which reflects abrupt neural shifts, respiratory flow evolves more gradually, making transitions appear less distinct and occasionally delaying recognition. Additional challenges occur in subjects with frequent and irregular stage shifts, where rapid transitions introduce complexities beyond what macro-respiratory signals can fully resolve. These findings highlight both the robustness and the limitations of our approach, pointing toward the need for refined modeling of REM-related features and improved handling of microstructural dynamics.

**Figure 5.**
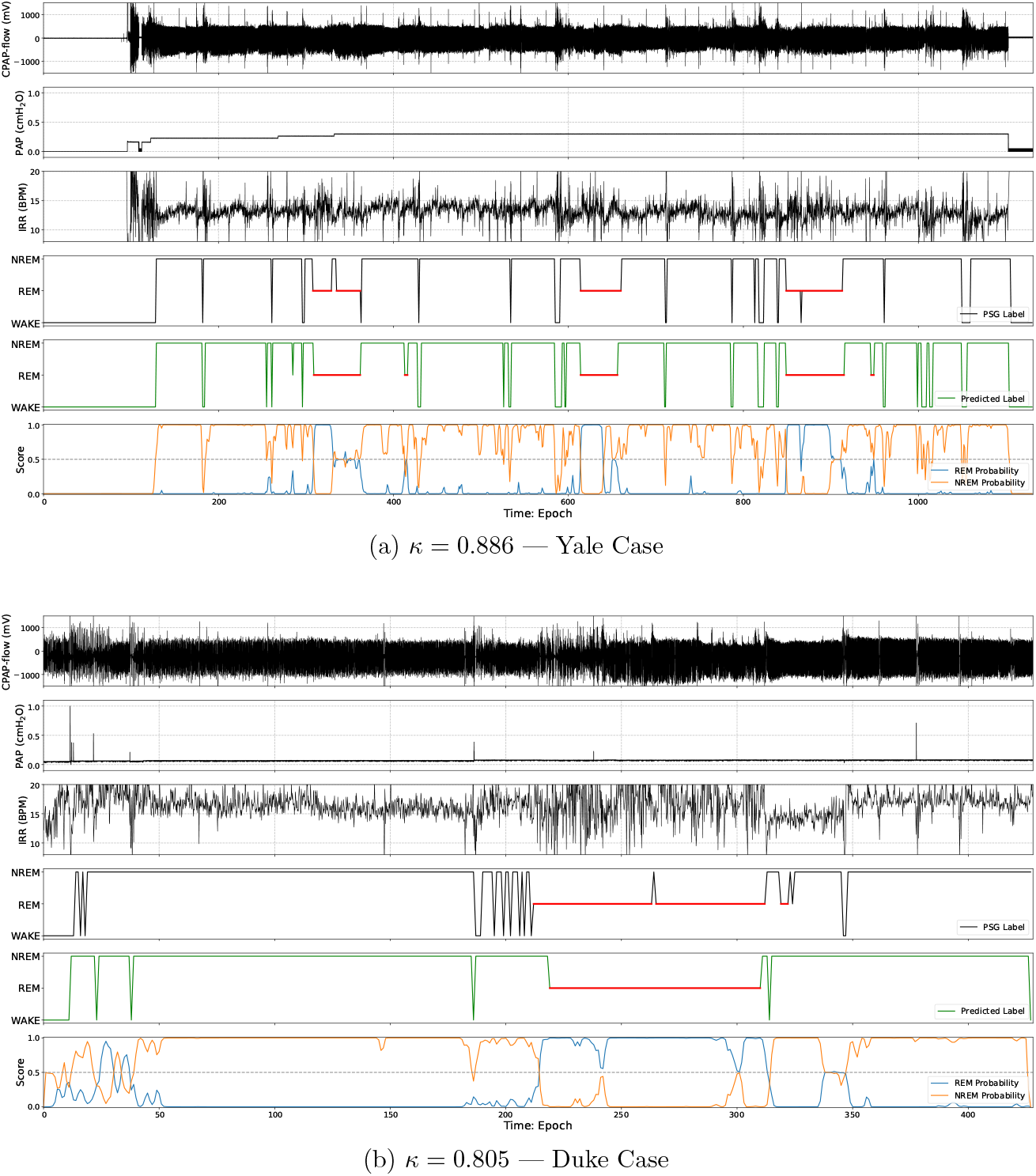
Illustration of results. Each panel shows representative overnight hypnograms with PSG labels, predicted labels, and stage probabilities.

**Figure 6.**
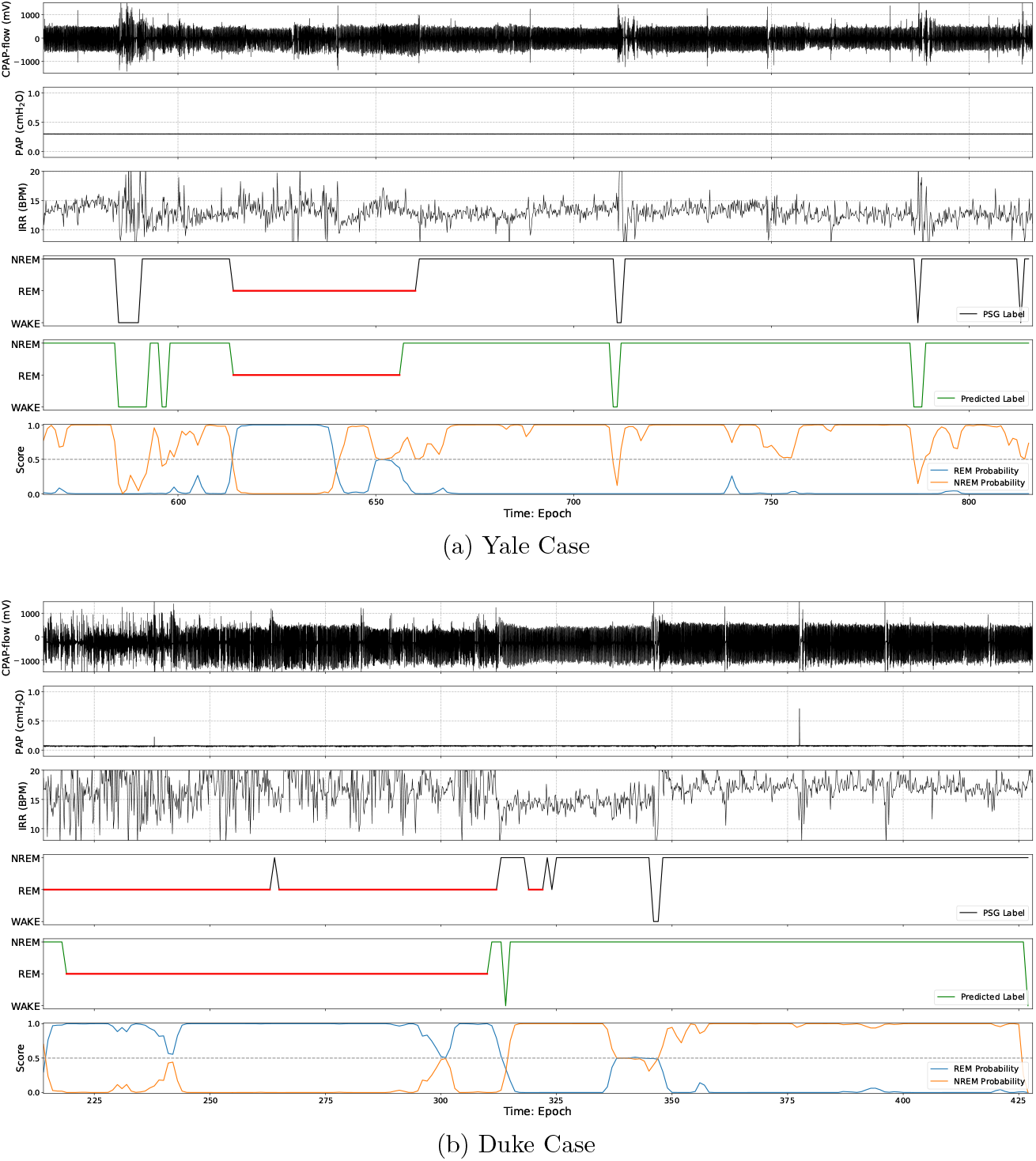
Zoomed-in plots of Figure 5.

### 4.4 Zero-shot Cross-dataset Transfer

To assess generalization under domain shift, we conducted zero-shot transfer [56] experiments by directly applying a model trained on Yale to Duke (denoted as Yale → Duke), and vice versa (denoted as Duke → Yale), without fine-tuning. As shown in Table 2, overall performance remained fairly stable across datasets, suggesting that the model captures sleep structure features not tied to a specific population or recording setup. The main weakness appeared in REM detection, where both sensitivity and precision dropped noticeably. This aligns with known clinical and technical challenges: REM is relatively rare, varies across individuals, and is more sensitive to device and scoring differences. In contrast, NREM staging transferred reliably, and Wake detection was mostly consistent, though with some asymmetry between transfer directions. Importantly, REM specificity remained high, indicating the model avoided over-predicting REM even if many epochs were missed.

**Table 2.** The zero-shot cross-dataset transfer results on Yale and Duke datasets.

| Duke → Yale | | | | | | | | | | | |
| --- | --- | --- | --- | --- | --- | --- | --- | --- | --- | --- | --- |
| Overall |  |  | Sensitivity |  |  | PPV |  |  | Specificity |  |  |
| Acc | $\kappa$ | F1 | Wake | REM | NREM | Wake | REM | NREM | Wake | REM | NREM |
| 69.7 | 0.459 | 0.627 | 0.722 | 0.449 | 0.738 | 0.601 | 0.464 | 0.803 | 0.827 | 0.925 | 0.718 |

| Yale → Duke | | | | | | | | | | | |
| --- | --- | --- | --- | --- | --- | --- | --- | --- | --- | --- | --- |
| Overall |  |  | Sensitivity |  |  | PPV |  |  | Specificity |  |  |
| Acc | $\kappa$ | F1 | Wake | REM | NREM | Wake | REM | NREM | Wake | REM | NREM |
| 68.9 | 0.432 | 0.616 | 0.595 | 0.414 | 0.810 | 0.577 | 0.570 | 0.755 | 0.872 | 0.930 | 0.620 |

A key difference emerged between transfer directions: Yale → Duke was more robust than Duke → Yale. This likely reflects the datasets’ characteristics. Yale comprises full-night titration studies with stable signal quality, enabling the model to learn clean respiratory patterns that remain valid under Duke’s noisier, split-night protocols. By contrast, Duke recordings are more fragmented due to sequential diagnostic and titration phases, frequent pressure changes, and movement, making them less effective as a training source for cleaner data. In short, models trained on clean, controlled data generalize more effectively to noisy environments than the reverse. This has practical implications: starting with a model trained on uniform datasets (like Yale) and lightly fine-tuning on heterogeneous targets (like Duke) is a more reliable deployment strategy than the opposite.

### 4.5 Demographic Subgroup Analysis

We further evaluated whether demographic factors affected model performance. Subjects were stratified by gender, BMI, and age, with cutoffs at BMI 30 [57] and age 60 years [58]. Subject-level *κ* scores were compared using the Mann–Whitney U test. As shown in Table 3, no statistically significant differences were found across any subgroup in either dataset (*p >* 0.05). At Yale, females achieved slightly higher agreement (*κ* = 0.614 vs. 0.592 for males) and older adults performed marginally worse (*κ* = 0.573 vs. 0.634 for *<*60 years), though not at a significant level (*p* = 0.056). At Duke, the gap between males and females was more pronounced (accuracy 77.9% vs. 69.1%), yet the small sample size (*N* = 16 per group) limited statistical power. BMI showed virtually no effect in either dataset (*p* = 0.183 at Yale, *p* = 0.983 at Duke). Taken together, these results suggest that the model generalizes robustly across demographic subgroups, with only mild, non-significant trends (e.g., younger subjects and females showing slightly higher performance). This indicates that demographic variability is unlikely to be the primary source of performance differences observed across datasets.

**Table 3.** Demographic subgroup analysis on Yale and Duke Datasets. Note that the p-value for each group was estimated using the Mann–Whitney U test based on the subject-level.

| Yale |  |  |  |  |  |  |
| --- | --- | --- | --- | --- | --- | --- |
| Group | N | Acc | $\kappa$ | F1 | p-value | Conclusion |
| Male | 47 | 77.3 | 0.592 | 0.716 | 0.579 | n.s. |
| Female | 45 | 79.4 | 0.614 | 0.735 |  |  |
| BMI < 30 | 33 | 76.8 | 0.574 | 0.703 | 0.183 | n.s. |
| BMI ≥ 30 | 59 | 79.2 | 0.618 | 0.737 | | |
| Age < 60 | 44 | 79.8 | 0.634 | 0.751 | 0.056 | n.s. |
| Age ≥ 60 | 48 | 77.0 | 0.573 | 0.699 | | |

| Duke |  |  |  |  |  |  |
| --- | --- | --- | --- | --- | --- | --- |
| Group | N | Acc | $\kappa$ | F1 | p-value | Conclusion |
| Male | 16 | 69.1 | 0.467 | 0.643 | 0.158 | n.s. |
| Female | 16 | 77.9 | 0.582 | 0.730 |  |  |
| BMI < 30 | 8 | 73.5 | 0.503 | 0.674 | 0.983 | n.s. |
| BMI ≥ 30 | 24 | 73.6 | 0.529 | 0.688 | | |
| Age < 60 | 21 | 74.4 | 0.531 | 0.692 | 0.267 | n.s. |
| Age ≥ 60 | 11 | 71.8 | 0.507 | 0.676 | | |

### 4.6 Impact of PAP

PAP stability plays a decisive role in classification accuracy, as shown in Table 4. To quantify PAP stability for each subject, we first normalized the PAP signal using min–max scaling [59]. We then computed the standard deviation (SD) of the normalized signal. Subjects with an SD greater than 0.2 were classified having low PAP stability, while those with an SD less than or equal to 0.2 were classified as having high PAP stability. At Yale, high-stability subjects achieved accuracy, *κ*, and macro-F1 of 79.3%, 0.617 and 0.744 respectively, compared to 77.6%, 0.593, and 0.708 respectively in low stability. Duke shows an even larger gap: high-stability subjects reached 77.0%, 0.606, and 0.743 respectively, while low-stability dropped to 72.4%, 0.493, and 0.664 respectively. The advantage is most evident in REM staging, where sensitivity improves from 0.553 to 0.650 at Yale and from 0.516 to 0.529 at Duke, while specificity remains consistently high (*>* 0.91). This pattern highlights that stable PAP suppresses baseline drift and irregular fluctuations in CPAP-flow, yielding clearer respiratory cycles and more distinguishable stage transitions. In contrast, frequent PAP adjustments distort airflow dynamics, masking subtle REM signatures and inflating inter-subject variability. The sharper decline in Duke suggests that PAP stability not only enhances within-subject signal clarity but also buffers against cross-subject heterogeneity, making the model more generalizable under diverse clinical settings.

**Table 4.** PAP feature analysis on Yale and Duke datasets: Comparison of performance between subjects with high and low PAP stability across both datasets.

| Yale |  |  |  |  |  |  |  |  |  |  |  |  |
| --- | --- | --- | --- | --- | --- | --- | --- | --- | --- | --- | --- | --- |
| PAP stability | Overall |  |  | Sensitivity |  |  | PPV |  |  | Specificity |  |  |
| | Acc | $\kappa$ | F1 | Wake | REM | NREM | Wake | REM | NREM | Wake | REM | NREM |
| High | 79.3 | 0.617 | 0.744 | 0.723 | 0.650 | 0.854 | 0.777 | 0.626 | 0.835 | 0.927 | 0.942 | 0.738 |
| Low | 77.6 | 0.593 | 0.708 | 0.774 | 0.553 | 0.822 | 0.770 | 0.488 | 0.846 | 0.916 | 0.918 | 0.768 |

| Duke |  |  |  |  |  |  |  |  |  |  |  |  |
| --- | --- | --- | --- | --- | --- | --- | --- | --- | --- | --- | --- | --- |
| PAP stability | Overall |  |  | Sensitivity |  |  | PPV |  |  | Specificity |  |  |
| | Acc | $\kappa$ | F1 | Wake | REM | NREM | Wake | REM | NREM | Wake | REM | NREM |
| High | 77.0 | 0.606 | 0.743 | 0.760 | 0.529 | 0.883 | 0.734 | 0.912 | 0.752 | 0.915 | 0.984 | 0.676 |
| Low | 72.4 | 0.493 | 0.664 | 0.670 | 0.516 | 0.799 | 0.671 | 0.537 | 0.791 | 0.905 | 0.913 | 0.665 |

### 4.7 Evaluating Potential Overestimation of CPAP Efficacy Using Mask-On Time

To assess the potential influence of CPAP mask-on time on sleep estimation and its clinical implications, we performed two additional analyses. The definition of mask-on time is when the mask is on the patient during the titration study (excluding bathroom breaks, mask-off times due to discomfort, etc). First, a Bland-Altman analysis in Figure 7 was conducted to compare PSG-derived mask-on TST with model-predicted mask-on TST, allowing us to quantify systematic bias and agreement between the two methods. Second, scatter plot analyses in Figure 8 were performed by plotting both PSG-derived and model-predicted mask-on TST against the total mask-on time, in order to evaluate the relationship between mask-wear duration and actual sleep time. As we know, when physicians monitor CPAP treatment, mask-on time alone is not sufficient, since what truly matters is how much of that time the patient is actually asleep. CPAP can only exert its therapeutic effect during sleep.

**Figure 7.**
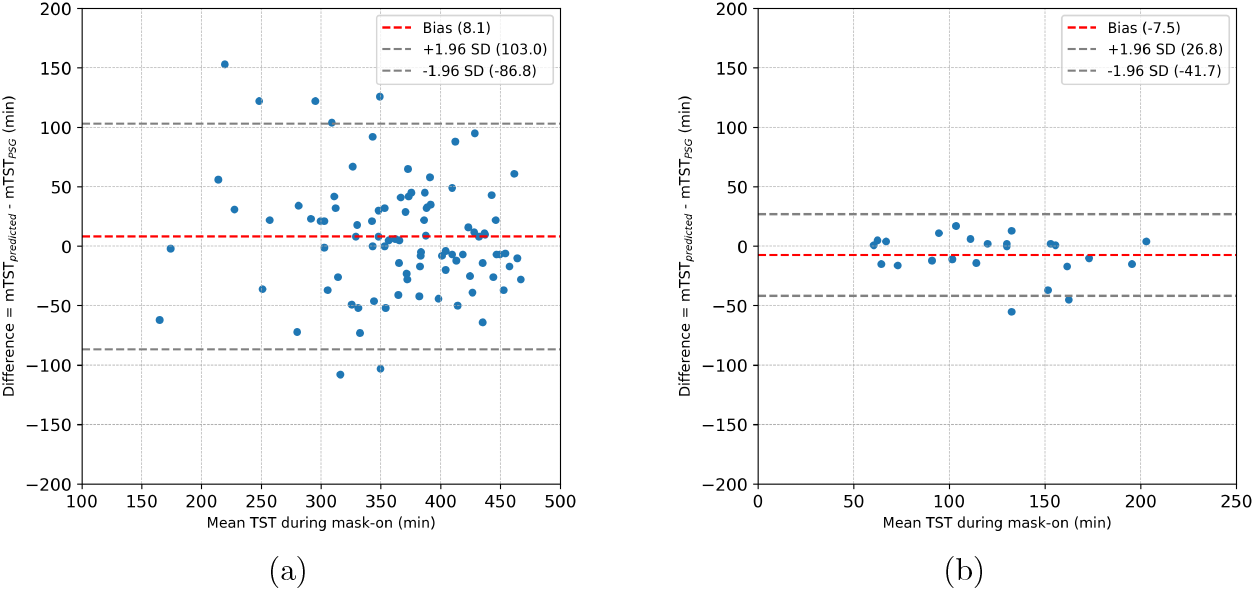
Bland–Altman analysis comparing mask-on predicted TST (mTST_predicted_) to mask-on PSG-TST (mTST_PSG_) on (a) Yale and (b) Duke datasets. SD: Standard Deviation (min: minute).

**Figure 8.**
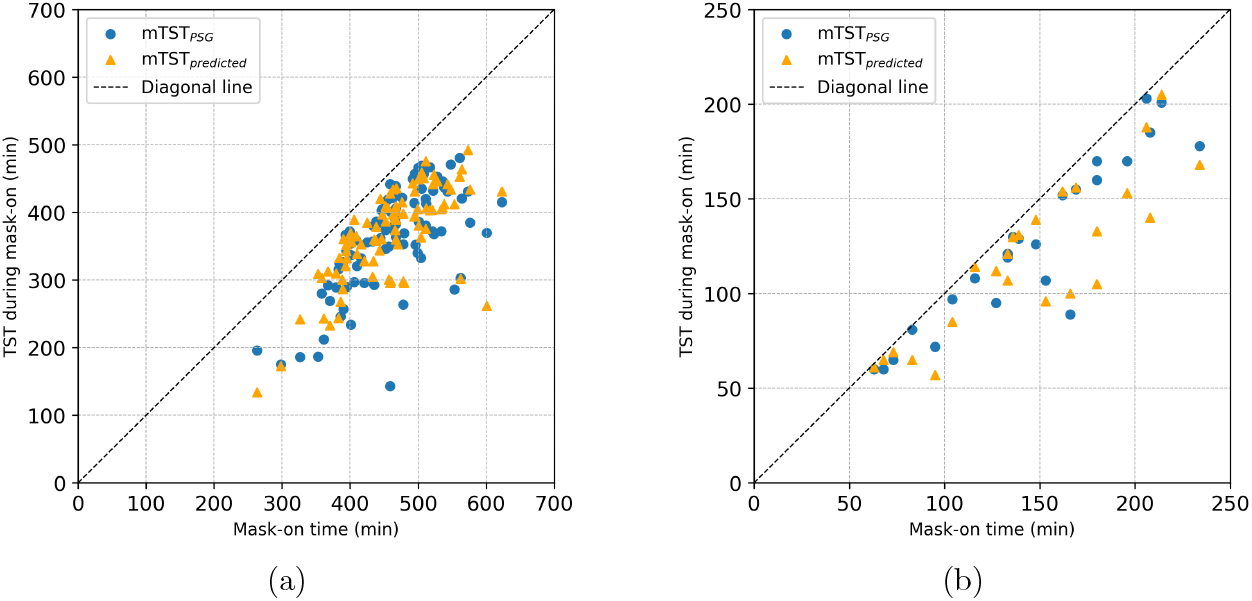
Scatter plot comparing mask-on predicted TST (mTST_predicted_) to mask-on PSG-TST (mTST_PSG_) on (a) Yale and (b) Duke datasets. (min: minute)

Therefore, providing a direct estimate of mask-on TST with our proposed method can make treatment follow-up more clinically meaningful. According to our results, although the model-predicted mask-on TST estimation still has room for improvement, it shows an acceptable level of agreement with PSG-derived mask-on TST.

### 4.8 Ablation Study

This section presents the ablation study, including both feature and fusion ablation, along with their implementation and corresponding results.

#### 4.8.1 Feature Ablation

Table 5 highlights the critical role of statistical feature augmentation in our framework. When window-level descriptive features, such as means, standard deviations, maxima, and minima, are excluded, the model relies solely on raw sample-wise representations. This simplification substantially degrades performance, with Cohen’s *κ* dropping by nearly 20% across both Yale and Duke datasets. Furthermore, sensitivity and PPV for REM classification decline by over 10 percentage points, reflecting the model’s reduced ability to capture subtle stage-specific variability. These results confirm that statistical aggregation provides complementary temporal context by summarizing local fluctuations and enhancing representation of stable breathing patterns. In particular, descriptive statistics mitigate redundancy in high-frequency samples while preserving discriminative power, thereby strengthening the model’s ability to distinguish between adjacent stages such as REM and light NREM.

**Table 5.** Ablation study comparing different feature augmentation sets (sample-wise versus statistic-wise) and fusion strategies (early fusion versus late fusion) on Yale and Duke datasets.

| Yale |  |  |  |  |  |  |  |  |  |
| --- | --- | --- | --- | --- | --- | --- | --- | --- | --- |
| Feature |  | Metric |  |  | Fusion |  | Metric |  |  |
| Sample | Statistic | Acc | $\kappa$ | F1 | Early | Late | Acc | $\kappa$ | F1 |
| v | x | 71.1 | 0.466 | 0.615 | v | x | 65.4 | 0.406 | 0.578 |
| x | v | 77.6 | 0.587 | 0.712 | x | v | 70.6 | 0.462 | 0.612 |
| v | v | 78.5 | 0.605 | 0.727 | v | v | 78.5 | 0.605 | 0.727 |

| Duke |  |  |  |  |  |  |  |  |  |
| --- | --- | --- | --- | --- | --- | --- | --- | --- | --- |
| Feature |  | Metric |  |  | Fusion |  | Metric |  |  |
| Sample | Statistic | Acc | $\kappa$ | F1 | Early | Late | Acc | $\kappa$ | F1 |
| v | x | 66.5 | 0.368 | 0.581 | v | x | 61.9 | 0.359 | 0.573 |
| x | v | 71.7 | 0.473 | 0.649 | x | v | 66.9 | 0.406 | 0.596 |
| v | v | 73.6 | 0.524 | 0.685 | v | v | 73.6 | 0.524 | 0.685 |

#### 4.8.2 Fusion Ablation

As shown in Table 5, the choice of fusion strategy has a decisive influence on classification accuracy. Early fusion, which directly concatenates feature channels at the input stage, yields the weakest baseline due to over-simplified interactions and the loss of modality-specific signatures. Late fusion, where each feature channel (CPAP-flow, PAP, IRR) is processed independently before integration, performs better by preserving modality-specific structure but fails to fully capture interdependencies. The strongest performance arises when combining both strategies in our dual-fusion design. This approach leverages the complementary strengths of early and late fusion: low-level features interact early to capture correlated dynamics, while modality-specific streams remain distinct until later integration, preserving channel-specific nuances. For instance, REM stage prediction on the Yale dataset improves markedly, with sensitivity and PPV reaching 0.604 and 0.558, representing a 50-70% gain compared to early (0.357/0.314) or late fusion (0.349/0.366). Similar patterns are observed on the Duke dataset, underscoring the robustness and generalizability of the combined approach.

## 5 Discussion

We propose a knowledge-informed DFMP-CNN framework that features the dual-fusion, multi-period design. Experimental studies in two databases collected from Yale New Haven Hospital and Duke University Hospital show its effectiveness in sleep staging, particularly capturing the subtle variability of sleep staging in CPAP-flow. Specifically, it shows promising potential to automatically score even the difficult-to-detect REM stage. The algorithm also shows its promising performance in estimating TST, a cornerstone metric in sleep medicine that directly impacts the validity of many downstream measures. For example, TST serves as the denominator for numerous derived indices such as the apnea–hypopnea index (AHI), sleep efficiency, and wake-after-sleep-onset (WASO). Errors in TST estimation can propagate through these metrics, potentially altering diagnostic thresholds and therapy evaluation. Moreover, because respiratory event burden is often stage-specific, reliable estimation of TST together with accurate REM/NREM distribution is essential for capturing REM-related AHI and other stage-dependent indices. By maintaining high fidelity in TST estimation even under the challenging conditions of CPAP titration studies, where frequent pressure adjustments fragment sleep, DFMP-CNN ensures that both absolute and derived clinical indices remain reliable and interpretable. This robustness has the potential not only to support diagnostic accuracy but also to enhance confidence in longitudinal monitoring and large-scale research applications where manual annotation is impractical.

In this study, we focus on classifying sleep stages into wake, REM, and NREM. Future research could extend its applicability to more granular sleep stage detection, particularly further classifying NREM into light sleep and deep sleep (slow-wave sleep, SWS). Detecting SWS remains challenging due to its relative scarcity in clinical datasets, particularly among older adults and patients with OSAHS. This scarcity leads to inherent class imbalance and limits the model’s opportunity to learn stage-specific features and generalize to new subjects. This limitation prevents a thorough exploration of SWS in the present study. We hypothesize that with access to a clinical database containing high-quality SWS labels, our model could generalize effectively to enable accurate automatic four-stage sleep classification from CPAP-flow. This hypothesis is based on respiratory dynamics during SWS. In SWS, breathing patterns are generally slower, more regular, and highly rhythmic. The CPAP-flow signal often exhibits longer and smoother waveforms with modestly reduced amplitudes, contrasting sharply with the irregular fluctuations of wakefulness or REM. Because these signals lack abrupt transitions, the model must focus on stable, low-amplitude features over extended temporal windows. This necessitates algorithms that are not only robust to noise and missing data but also capable of integrating long-range temporal dependencies. We expect to explore this possibility in our future work.

Clinically, the implications of this work extend beyond automatic epoch-by-epoch stage classification. Prior studies have shown that incorporating sleep to wake transitions can guide therapy optimization [6, 7]. Building on this, our model has the potential to enable personalized, stage-adaptive pressure modulation in CPAP or auto-adjusting positive airway pressure (APAP) systems. For example, higher sensitivity during REM could counteract increased upper-airway collapsibility, while reduced pressure during stable NREM or SWS could minimize patient discomfort.

Such adaptive strategies have the potential to improve therapeutic effectiveness, enhance long-term adherence, and advance patient-centered care.

This study has several limitations. First, the sample size is modest and restricted to subjects suspected or diagnosed with OSAHS, and the sleep labels are inherently imbalanced. While the inherent label imbalance might be challenging to improve, expanding and diversifying the dataset is possible and will be critical. A larger corpus with balanced representation across all sleep stages would enhance the model’s ability to capture inter-stage boundaries, reducing confusion between adjacent stages such as REM and NREM. Second, it is well recognized that deep learning models operate largely as “black boxes”, with limited theoretical justification beyond simplified settings [17, 18, 60]. Although our model was developed with qualitative physiological guidance and validated through multiple analyses to mitigate overfitting, its underlying mechanisms remain poorly understood quantitatively. Advancing theoretical frameworks to elucidate how such models function could not only deepen our understanding but also inform improved model design.

## 6 Conclusion

We developed DFMP-CNN, a robust framework that effectively captures multi-scale temporal patterns corresponding to distinct sleep stages, for sleep stage classification using CPAP-flow. Experimental results confirm its ability to detect clinically meaningful sleep stages and transitions. Overall, this study lays the groundwork for reliable CPAP-integrated sleep monitoring and analysis in both clinical and home environments.

## Data Availability

The data that support the findings of this study are available from the corresponding author upon reasonable request. These datasets are not publicly available due to restrictions from the institutional review boards of Yale University School of Medicine and Duke University Hospital.

## Acknowledgements

The authors would like to thank the staff and collaborators at PranaQ Pte. Ltd., Yale New Haven Hospital, Duke University Hospital, and New York University for their support and valuable discussions.

